# Empowering Women in Healthcare: Unveiling Their Experiences and Strategies for Organizational Support

**DOI:** 10.1101/2023.10.06.23296671

**Authors:** Abi Sriharan, Nigar Sekercioglu, Whitney Berta, Sylvain Boet, Audrey Laporte, Gillian Strudwick, Senthujan Senkaiahliyan, Savithiri Ratnapalan

**Affiliations:** York University and University of Toronto; McMaster University; University of Toronto; University of Ottawa; Centre for Addiction and Mental Health and University of Toronto; M.Mgt.AI, University of Toronto; Hospital for Sick Children and University of Toronto

**Author notes:** **Corresponding author’s complete contact information:** Abi Sriharan, MSc, DPhil, Krembil Centre for Health Management and Leadership, Schulich School of Business, York University, Toronto, Canada. **Funding:** This research is funded through a grant from the Canadian Institute for Health Research Catalyst Grant - Quadruple Aim and Equity, RN474112 - 475348.

**Keywords:** healthcare workforce, women, retention, talent management, health human resources

## Abstract

**Importance:** Health care systems worldwide are grappling with rising burnout among health care workers, leading to increased rates of early retirement and job transitions. This crisis is detrimentally affecting the quality of patient care, contributing to long wait times, decreased patient satisfaction, and a heightened frequency of patient safety incidents and medical errors. Notably, women constitute 70% of the health care workforce.

**Objective:** The primary objective of this study is to uncover the factors influencing the turnover intentions and sustained commitment of HCWs who self-identify as women.

**Design, Setting, and Participants:** We used grounded theory in this qualitative study. From January 2023 to May 2023, we conducted individual semi-structured interviews with 27 frontline HCWs working in Canada and representing diverse backgrounds. The data underwent thematic analysis, which involved identifying and comprehending recurring patterns across the information to elucidate emerging themes.

**Results:** In the analysis we uncovered three factors influencing women’s intent to exit the frontline workforce: organizational, professional, and personal. Organizational factors related to work related policies, compensation, positive work culture, and effective leadership behaviors emerged as essential elements for retaining women in health care organizations.

**Conclusions and Relevance:** The outcomes of this study shed light that women’s intention to leave frontline clinical roles is shaped by three interacting factors: personal, professional, and organizational. Although the personal factors are beyond the scope of organizations in retaining women in the frontline clinical care, organizations can shape organizational strategies, organizational culture and leadership approaches to ensure they are women friendly and transform the organizational environment by creating a thriving culture for women to perform their professional role in the organizations within the constraints of their personal circumstances, such as care giving responsibilities at home.

**Key Points:** *Question:* Why do women in health care depart from frontline clinical practice, and what proactive measures can organizations implement to ensure their continued presence and contribution to patient care at the forefront?

*Findings:* In this qualitative study, involving interviews with a diverse group of health care professionals who self-identify as women, participants pinpointed three interconnected factors influencing their choices to exit clinical practice: personal circumstances, professional roles, and the organizational context. They emphasized that fostering an organizational culture that supports women, offers equitable rewards, and provides robust and supportive leadership is imperative for retaining them in frontline positions.

*Meaning:* Although personal circumstances and the inherent nature of professional roles may be beyond the direct control of organizations, they can actively shape the organizational context to create a more women-friendly environment. This reshaping entails fostering a supportive organizational culture for women, implementing fair and equitable reward systems, and providing comprehensive training for managers and leaders in talent management strategies. These concerted efforts can significantly contribute to retaining women within frontline work environments.

---

Global health systems are facing a significant workforce crisis due to increasing levels of burnout, early retirement, and workforce transitions among health care professionals, which is having serious implications for health care quality and patient outcomes (Casey, 2023; Cutler, 2022; Linzer et al., 2022; Poindexter, 2022). The consequences of this crisis are far-reaching, including long patient wait times, low patient satisfaction, and increasing patient safety incidents and medical errors (Canadian Medical Association, 2022; Melnick et al., 2023).

In general, job demands, work conditions, work relationships, and organizational culture are considered essential elements for job satisfaction and work–life balance (Adams et al., 2021; de Vries et al., 2023; Steinmetz et al., 2014; Wakerman et al., 2019). When these factors erode, employees leave their organizations.

However, the experiences of women are unique. Women in the health care industry, similar to their counterparts in other sectors, often have to balance their professional and personal lives. However, gender bias remains a persistent problem in health care (Hui et al., 2020; Salles et al., 2019). This bias is exacerbated by the demanding nature of the health care profession, which involves long hours, including after-hours and weekend work; emotional labor; and a relentless pace (Sriharan et al., 2020; Sriharan, Ratnapalan, et al., 2021; Sriharan, West, et al., 2021).

Women make up 70–80% of health care workforce globally (Sriharan et al., 2021). As the world slowly recovers from the recent COVID-19 pandemic, health care systems have to increase services to deal with the backlog of postponed services in addition to providing new and ongoing services to the public (Bagenal, 2022; Ghoshal et al., 2022; Walker et al., 2022). To address the surge in health care service needs and to compensate for the health care workers (HCWs) who left the frontline clinical care, especially during the COVID-19 pandemic, health care organizations and health systems are currently engaged in significant recruitment to meet health care workforce demand. (Frogner & Dill, 2022; Iacobucci, 2021; Waters, 2022; Wilkinson, 2023). However, recruitment efforts alone are not sustainable. It is equally important to ensure that the current workforce, as well as the newly recruited clinical staff, feels supported to stay in the workforce (Walker et al., 2022). This study aims to explore the reasons women HCWs leave frontline clinical care and to identify ways organizations can support them to continue in the workplace.

## Methods

### Design

We undertook an explorative grounded theory study (Charmaz, 2014; Strauss & Corbin, 1997) to understand how women HCWs experience work and explore what employers can do to support them at work.

### Setting and Sample

The study took place in the province of Ontario in Canada. Canada’s health care system is primarily funded by the government, with around 70% of its financing coming from public sources. This publicly funded system ensures that all residents have access to essential medical services. Most physicians work as independent contractors in health care facilities and receive payments through a single-payer system. A wide range of health care professionals, including nurses and allied health professionals, are employed by hospitals, clinics, long-term care facilities, and home health care agencies to provide comprehensive care to the population. These organizations are publicly funded but privately delivered nonprofit independent corporations. We employed a purposive sampling method to recruit women in health professions actively engaged in frontline clinical care roles in Ontario.

### Recruitment and Participants

We established specific criteria for participant inclusion in our recruitment process: (a) self-identifies as a woman; (b) is a health care professional in Ontario, which includes licensed physicians, registered nurses, registered practice nurses, nurse practitioners, and other health care providers such as personal support workers and physician assistants; and (c) has engaged in direct patient care within a health care delivery setting, either currently or within the past 24 months. We targeted recruitment to ensure participation from individuals with diverse identities. Our recruitment strategy encompassed email campaigns, social media posts, Listservs, and recruitment flyers. Participants who completed the intake form online and provided online informed consent, were contacted by the research team for an online interview. During the qualitative interviews, we continued our recruitment efforts until we reached a point of data saturation (Saunders et al., 2018)—that is, where no new substantial insights were emerging.

### Data Collection

We implemented a semi-structured one-to-one interview process. During the interviews, we allowed participants to reflect upon their experiences as HCWs. The primary interview inquiries concerned participants’ experiences, expectations, and intentions regarding their current and future careers.

Before the interviews, we emailed informed consent forms to the invited participants. Ensuring strict confidentiality, we clarified that participants had the right to withdraw from the interview at any point. We treated the data we obtained from each interview with confidentiality and stored them securely in a protected database. We conducted the interviews via Zoom to maintain consistency, with one interviewer trained in the interview guide conducting all interviews. We conducted the interviews exclusively in English to maintain uniformity.

The interview guide we used and relevant materials are available from the Open Science Framework. All participants were invited to complete an optional structured personal data form, which gathered information about demographics (age and cultural background) and lifestyle details (current employment status, annual income, health care work duration, and parenting status).

### Ethics

The study commenced after ethics approval from the University of Toronto’s Research Ethics Board (REB 43932). As part of our recruitment process, we gave participants an electronic informed consent form to fill out. Once they completed the form, our research assistant scheduled virtual interviews. Participants were given $60 gift cards as a token of appreciation for completing the interview.

### Data Analysis

Interviews were recorded, professionally transcribed verbatim using MAXQDA transcription tool, deidentified, manually cleaned and reviewed for accuracy. Transcripts were not shared with the participants. Using grounded theory, we conducted open coding on emerging data from the interviews. In the open coding process, we systematically divided the data into smaller segments and assigned descriptive codes to them. This approach allowed us to explore various ideas and concepts within the data without any preconceived notions.

Once all interview transcripts were coded, reports were generated that collected all the quotations assigned to the same code. We then used a thematic analysis approach to identify and analyze repeated patterns across data and explain emerging themes (Boyatzis, 1998). First, through an iterative process, the code reports were examined together to develop axial codes. We used axial coding to establish connections and relationships between the initial codes, and we organized them into broader themes based on our guiding research questions. We then analyzed how the themes are related to each other to identify the core concepts that underlie the data. We kept detailed notes of the decisions, questions, and comments.

To increase the rigor of our analysis, two independent researchers (NS, SS) coded all the data. A third researcher (AS) reviewed all the codes, themes, concepts, and quotes, and then three researchers (AS, NS, SS) met to resolve any discrepancies.

## Results

We included 27 participants in this study (nine physicians [33.3%], 15 nurses [55.5%], and three [11.2%] allied health care professionals). We received 25 responses to the demographic survey. Participants represented diverse age, racial, experience, and income backgrounds (Table 1).

### Factors Affecting Women’s Intentions to Leave Direct Patient Care Roles

Regardless of the participants’ professional backgrounds, we noticed that similar themes started to emerge during the interviews about women’s experiences in the workplace and the significant factors affecting their intentions to leave frontline clinical work. We grouped the experiences into three categories: organizational factors, professional factors, and personal factors. Table 2 provides a summary of themes and illustrative quotations.

### Organizational Factors

Participants emphasized how factors within the organization played a crucial role in their decisions to move away from direct patient care roles. Workload emerged as a major theme of concern for the participants. Although workload issues were significant during the COVID-19 pandemic, these issues have persisted in the post-pandemic period as well. Most of the participants highlighted that existing staff are exxperiencing increased workload due to significant staffing shortages in many healthcare organizations., the. Many expressed the feeling that they were not fairly compensated for their increased workload. Further, the increased work often meant their work conditions changed and required them to work longer hours, which made them feel emotionally drained as it took away their time outside of work. Participants also felt the existence of biases related to gender and race played a role in compensation differences, some individuals were favored by the organizations over others in terms of getting opportunities for advancement and better paying roles. Individuals voiced their concerns about organizational leadership frequently neglecting to consider the distinctive experiences of women, often imposing work schedules without prior consultations. Many shared the sentiment that in environments lacking supportive colleagues and empathetic leaders and managers capable of comprehending the unique challenges faced by women, individuals are more inclined to depart from frontline clinical care.

### Professional Factors

Participants stated that they became aware of the expectations of their professional roles and the limitations of the expectations during the COVID-19 pandemic. For example, participants who worked directly with patients had to stay at the health care facilities while providing direct patient care. To make the situation more complex, there were restrictions on how care could be given and the resources available during COVID-19. As a result, there often needed to be a better match between what was considered the best care based on clinical judgment or patient expectation and what the health care system could practically provide.

Further, the participants perceived the hierarchy of clinical medicine as a concern. For example, one participant mentioned that when their colleagues, specifically senior physicians and administrative staff, were working remotely and that people providing in person patient care not only dealt with medical issues of the patients but also attended virtual meetings, often involving discussions related to resources and how care was given. Although participants highlighted the purpose and passion they have for clinical work and how it motivated them to continue in the face of adversity, they questioned the meaning of their purpose when organizational circumstances such as workload, gender bias, work conditions, and work environment were not conducive for their optimal functioning.

### Personal Factors

Most participants highlighted they value the importance of work–life balance. Many participants grappled with balancing their workload with personal responsibilities, including caring for their families, which raised doubts about the sustainability of their current work situations. Participants highlighted that women have unique challenges not only at work but also at home, and when they are faced with a work situation that impacts their personal life, they often tend to leave their professional role. A sense of fairness emerged as an important motivation that drives the individuals to feel supported to stay in the workforce. Since the onset of the pandemic, participants have experienced heightened concerns about their safety.

Witnessing their colleagues and patients contract COVID-19 and, tragically, succumb to it or live with life altering sequale, led to their experiencing a deep-seated sense of trauma.

Additionally, some participants shared that the emotional turmoil patients and their families experienced seemed to spread, affecting their own stress and anxiety levels. Furthermore, the departure of coworkers for improved prospects elsewhere evoked a desire among some participants to explore alternative opportunities.

### Resources to Support and Retain Women in Health Care

Participants identified the importance of organizational strategy, organizational culture, and leadership support as essential factors that can help shape organizational approaches to retain women HCWs in frontline clinical roles. Table 3 provides a summary of themes and illustrative quotations.

### Organizational Strategy

Organizational strategy includes elements of organizational elements-related workforce that organizations can invest in modifying to ensure their organization is women friendly.

Participants emphasized the significance of fair and equal compensation and benefits, such as paid leave and access to free medical services, as pivotal factors that motivate them to stay in the workforce. There was a perception that women are not paid equally for their work efforts. To retain them in the workforce, organizations should ensure fair compensation, respectful work conditions, equitable benefits, and policies that are inclusive of women’s unique circumstances. In addition to the compensation and benefits, participants identified the importance of organizational investment in their personal and professional growth through professional development training that could substantially enhance their skill sets and their opportunities for advancement. Participants also highlighted that the virtual work options available during the COVID-19 pandemic provided a positive experience for many women HCWs. Organizations should consider investing in resources to integrate virtual work opportunities when feasible for women HCWs, especially in situations such as administrative work and remote monitoring.

### Organizational Culture

Participants frequently voiced a need to be adequately recognized for their contributions and respected for their skills, especially in team-based settings where unspoken professional hierarchies exist. Further, participants emphasized the necessity of addressing biases about the gender and racial backgrounds of the professionals to ensure the work environment is fair and inclusive.

They underscored the critical importance of fostering a workplace culture that appreciates individuals’ contributions in team-based care, the importance of being treated respectfully, and having their capabilities trusted by team members. Additionally, participants emphasized the importance of open communication of concerns and facilitating shared decision-making regarding patient care decisions. Further, participants identified the importance of ensuring that organizations create a supportive atmosphere for women, regardless of their racial or professional role, to share their ideas, engage in the decision-making process, and obtain opportunities for professional advancement.

### Leadership

Participants consistently pointed out that microaggressions by and implicit behaviors of team leaders and other health care professionals shaped less favorable work experiences.

Participants mentioned that leaders have a crucial role to play in cultivating a supportive working condition where they are receiving constructive feedback for professional growth. Further, leaders’ ability to empathize with women’s unique experiences will improve work conditions and will help individuals to sustain their tenure in the clinical roles.

## Discussion

In the ever-evolving health care landscape, talent management has become a pivotal concern for organizations aiming to attract, nurture, and retain their most prized assets—their employees. We delved into the reasons that often drive women in the health care sector to exit the workforce, offering strategies organizations can adopt to preserve their female talent.

In our study we illuminated the multifaceted nature of the turnover rate for women HCWs, shaped by individual, professional, and organizational dynamics. It highlighted the importance of organizational strategies, organizational culture and leadership in supporting women HCWs to sustain in the frontline clinical care.

Women working in health care often face stereotypes and prejudices that suggest they cannot balance long hours, irregular shifts, and high-stress environments with family life. However, our research challenges this assumption and shows that the reasons for women leaving frontline health care are multifaceted.

The findings of this study indicate that women’s desire to leave frontline clinical roles is influenced by three interrelated factors: personal, professional, and organizational. Although personal factors are beyond the control of organizations when it comes to retaining women in frontline clinical care, organizations can invest in organizational strategies and culture and leadership by creating an environment that supports women, thereby helping them to perform their professional roles while balancing their personal responsibilities. This finding aligns with turnover theory, which postulates employees who manifest elevated levels of job satisfaction, maintain a harmonious equilibrium between work and personal life, exhibit an unwavering commitment to their organizations, and receive substantial support from their management are distinctly less predisposed to entertain thoughts of relinquishing their current employment positions (Hom et al., 2017).

Similarly, our findings align with the social exchange theory, which underscores that employees who receive favorable treatment, support, and recognition from their organizations are more likely to feel obligated and loyal to them, thereby boosting retention rates (Cropanzano & Mitchell, 2005).

By employing a qualitative approach, we unearthed valuable insights into the catalysts underlying individuals’ intentions to depart from frontline work. Future research endeavors focused on talent retention should draw insights from the emerging evidence we have generated in this study. These insights can serve as a foundation for prospective quantitative investigations. Such endeavors can uncover correlations between personal attributes, professional considerations, and organizational conditions and the intent to leave.

## Implications to Practice

Traditionally, within health care organizations, talent management tasks such as compensation, benefits, and training fell under the purview of specialized human resource professionals. However, in our study we underscored the pivotal role leadership and culture, in addition to organizational strategy, play in fostering effective talent management. Organizations should embrace an organization-wide approach to address workforce retention strategies beyond conventional human resources boundaries. Organizations should also equip their leaders and managers with the requisite skills to manage and retain their invaluable workforce effectively.

Furthermore, with the power of AI and data analytics, organizations can utilize people analytics capacity to conduct annual organization-centric analyses to contextualize the findings from this study. Measuring the experiences of HCWs working in the organization and formulating potent retention strategies specific to the organization or unit will foster an environment that nurtures job contentment, allegiance to the organization, and avenues for growth.

## Conclusion

Health care organizations must invest in resources that transcend conventional HR paradigms, cultivating engagement and motivation among health care professionals. Working in frontline clinical care presents a range of challenges for women. It is the responsibility of organizations to develop work environments that support women health care workers. This goal can be achieved by reviewing the work rewards systems, organizational culture, and leadership behaviors within the organization. By doing so, women will not only feel supported but also empowered to excel in their professional careers.

## Data Availability

The interview transcripts are saved in a secure database as required by ethics.

## Acknowledgement

Authors acknowledge Samah Hassan for her assistance with conducting interviews.

